# Structural, Behavioral, and Symptomatic Predictors of Risk Allele Frequency at rs10866912

**DOI:** 10.1101/2024.12.04.24318503

**Authors:** Katherine L. Forthman, Sathish Periyasamy, Rayus Kuplicki, Martin P. Paulus, Bryan J. Mowry

## Abstract

This study examines the distribution of risk (A) alleles at rs10866912 among 480 individuals stratified by group assignment, gender, age, race/ethnicity, body mass index (BMI), and education level. Preliminary analysis indicated higher prevalence of the risk allele in the population diagnosed with a psychiatric disorder, in concordance with past research. Linear models were applied to elucidate the relationship between the risk allele and various measures covering drug use, diet, psychiatric symptoms, neuropsychiatric ability, and structural brain measures. The findings highlight significant variations in allele frequencies across these measures, in particular egg consumption and analgesic use, which may provide insights into the mechanisms by which this genetic marker influences development of psychiatric disorders.

## Introduction

The single nucleotide polymorphism (SNP) rs10866912 has been implicated in schizophrenia based on a genome-wide association study (GWAS). Specifically, the single nucleotide polymorphism (SNP), rs10866912, reached genome-wide significance in the first published Indian-ancestry genome-wide association study (GWAS) of schizophrenia (1). The risk allele of this SNP was observed to downregulate NAPRT1, a gene that codes for the rate-limiting enzyme in the Preiss-Handler pathway in niacin metabolism. This pathway is believed to be the main source for nicotinamide adenine dinucleotide (NAD+) biosynthesis in the brain. NAD+ is a coenzyme essential for many cellular processes, including energy metabolism, DNA repair, and cell signaling. Given the brain’s high energy consumption and the vulnerability of neurons to metabolic stress, the efficient synthesis of NAD+ through the Preiss-Handler pathway has been hypothesized to have profound implications for cognitive function and the overall maintenance of neural health. We sought to assess the relationship of this SNP across various behavioral and structural measures. This detailed analysis explores the association of rs10866912 and drug use, diet, psychiatric symptoms, neuropsychiatric ability, and structural brain measures including cortical and subcortical volumes, cortical thickness, and sulcal depth within a cohort of 500 participants from the T1000 study and discusses its potential functional roles.

### Genetic AND Functional Context

The rs10866912 SNP is located in an intergenic region on chromosome 8q21.13 that may influence gene regulation. SNPs in regulatory regions can affect gene expression, contributing to phenotypic diversity and disease susceptibility. Trait-associated SNPs, including rs10866912, are often enriched in genic and regulatory regions such as enhancers and promoters, critical for transcription regulation (BMC Genomics) (<u>BioMed Central</u>).

### Association with Diseases

Several studies have identified rs10866912 as a potential risk factor for various health conditions. Here, we highlight its associations with specific diseases.

The SNP rs10866912 is associated with psychiatric disorders including mood and anxiety disorders and substance use disorders. Studies have shown a higher frequency of the risk (A) allele in individuals with mood and anxiety disorders compared to healthy controls, suggesting it may influence the genetic predisposition to these disorders (2). The risk allele is also more prevalent in individuals with substance use disorders, indicating that rs10866912 may play a role in the neurobiological pathways underlying addiction and substance use behaviors (3). The SNP is further associated with metabolic conditions, with previous research indicating a link between the SNP and obesity and BMI traits, suggesting that this variant may influence metabolism, affect energy balance and fat storage and contributing to obesity risk (4). Finally, rs10866912’s role in regulating genes involved with inflammation and metabolism suggests a possible link between the SNP and cardiovascular disease, though further research is needed to clarify the association. Previous research indicates rs10866912 as having impacts across a breadth of health symptoms, indicating that it may play a mechanistic role in the development of these traits. Incorporating rs10866912 into predictive models, such as linear regressions, can enhance our understanding of the biological pathways through which this SNP influences negative health outcomes.

## Methods

### Cohort

Our cohort included the first 500 participants in the Tulsa 1000 study (T1000) (5). Participants provided written informed consent and received remuneration for study participation according to the study protocol approved by the Western Institutional Review Board. The T1000 initiative collected a variety of longitudinal data on 1000 participants in Tulsa, Oklahoma. This study included healthy controls as well as individuals with mental health disorders including substance use disorder, depression, anxiety, and eating disorders. Data collected included basic demographics, behavioral data, mental disorder symptom questionnaires, and genetic data. Our study included only the first 500 participants to allow confirmatory analysis in the second half of the population in a future paper. 20 participants were removed from the dataset because they were missing the outcome variable.

### Genotyping and imputation

Genotyping was performed on participant blood samples using the Illumina Infinium Global Screening Array-24 (v.2.0) BeadChip arrays by RUDCR Infinite Biologics. Imputation was performed using the Michigan Imputation Server Pipeline (Minimac4, version 1.2.4) using the HRC reference panel (6).

Genetic information was imputed from 569,641 to 40,359,612 SNPs. Within the imputed dataset, we identified the SNP rs10866912

### Linear comparisons

We performed linear tests in order to determine relationships between the number of risk alleles and various predictors. Given the broad possible impact of the SNP, we investigated its effect on (1) psychiatric symptoms, (2) structural brain characteristics, and (3) lifestyle and nutritional variables. Specifically, we examined 331 predictors from the following categories: symptoms (34), volume (68), subcortical volume (41), thickness (68), sulcal depth (68), neuropsychiatric (40), drugs (6), and diet (6). This comprehensive selection of variables aims to capture the complex interplay between genetic predisposition and various neurobiological and behavioral endophenotypes. Notably, we include brain structure-related endophenotypes (volume, subcortical volume, thickness, sulcal depth) and neuropsychiatric markers that have been robustly associated with symptomatology (7), reflecting the underlying neural mechanisms potentially influenced by rs10866912. In addition, drug use is considered as a potential confounder due to its known impact on brain and behavior (8, 9). Dietary factors, particularly in the context of our niacin hypothesis, are included as key variables of interest, because genetic variation can significantly influence dietary choices (10). This approach allows us to explore how rs10866912 might contribute to the observed phenotypic variability, considering both direct effects on brain structure and function and indirect influences mediated by lifestyle factors. Covariates in the model included age, gender, education, BMI, and cortical volume. A separate regression was performed for each predictor. Predictors were normalized using optLog (11) prior to fitting. Variable importance was calculated as the difference in model R2 of the linear model with and without the predictor variable (i.e. the covariates only model). FDR and Bonferroni corrected significance were both computed to elucidate significance after adjustment for multiple comparisons.

## Results

From the 500 participants included in this study, we were able to identify the allele in 480 of the participants. Most of the population had 1 copy of the allele (*N* = 225, 47%). Within the remaining participants, 128 had 2 copies of the allele (27%), and 127 had no copies of the allele (26%). Of the Healthy Controls, a majority had no risk alleles (49.1%), whereas in the disordered populations the majority had either 1 or 2 alleles (Mood/Anxiety, 1 allele, 49.2% | substance use disorder, 1 allele, 49.3% | eating disorder, 2 alleles, 40.7%). Males and Females had a similar proportion of individuals with no risk alleles (Male, 26.6% | Female, 26.4%), however females more often had 1 allele than males (Male, 43.8% | Female, 48.6%). Among the different age groups, an absence of risk alleles was most common in the 20-30 group (28.7%) and least common in the 50-60 group (21.4%), 1 risk allele was most common in the 15-20 group (59.1%) and least common in the 30-40 group (44.7%), and 2 risk alleles was the most common in the 30-40 group (30.3%) and least common in the 15-20 group (13.6%). Stark differences were observed in allele count amongst different racial/ethnic categories. Having 1 allele was most common in white, Native American, and Hispanic participants (47.8%, 50.6%, and 63.2%, respectively). Within the Black population, all participants had at least one copy of the risk allele, with a majority having 2 copies (74.2%). BMI was observed to be slightly higher in the population with no risk alleles (28.7), followed by the population with 2 copies of the risk allele (28). Across all education levels, the majority had 1 risk allele.

Linear models identified 33 variables that significantly predicted risk allele count after controlling for the covariates age, gender, education, BMI, and cortical volume (p-value < 0.05). None of these findings survived either FDR or Bonferroni correction for multiple comparisons. Significant symptom variables included measures from the PROMIS bank (applied cognitive ability, ability to participate in social activities, sleep impairment, depression, activity satisfaction, anxiety, and informational support), and the PHQ sum score. Among the neuropsychiatric variables, significant relationships were discovered within the DKEFS Color Word (CW) and Verbal Fluency (VF) tests. These included from the CW task Inhibition/Switching Total Uncorrected Errors and Cumulative Naming Errors, and from the VF task Repetition Errors. Within the drug and diet categories, analgesics and eggs in diet were significant. Significant volumes included cortical volume right caudal anterior cingulate, cortical volume right frontal pole, and cortical volume left lateral occipital. Significant cortical thicknesses included left and right supramarginal, left and right transverse temporal, left isthmus cingulate, left pericalcarine, left precentral, left rostral anterior cingulate, right superior frontal, and right temporal pole. Significant sulcal depths included right lateral occipital, right caudal middle frontal, and left caudal anterior cingulate. Finally, significant subcortical volumes included optic chiasm, left ventral dc, brain stem, and left putamen (Figure 1).

**FIGURE 1.**
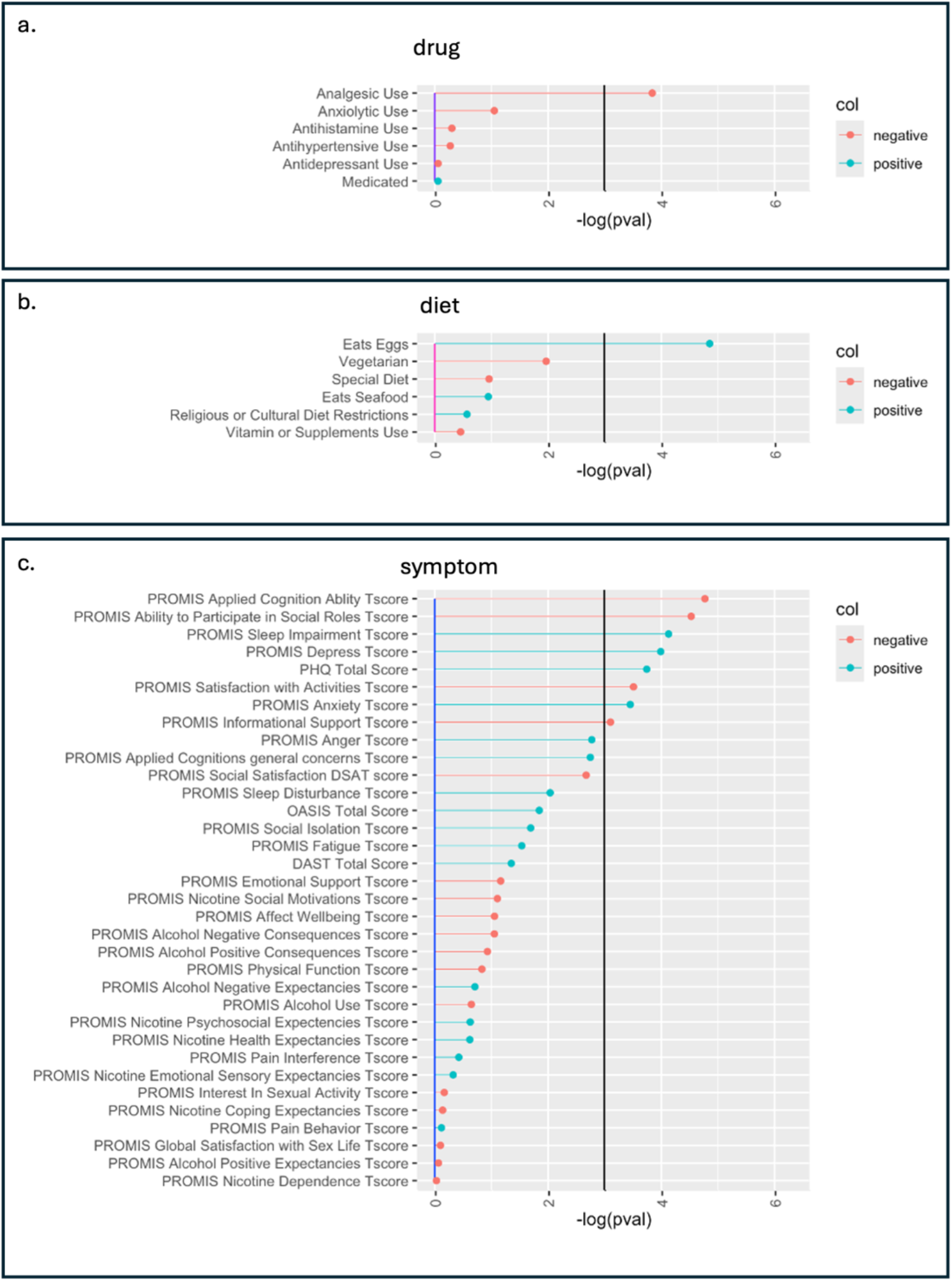
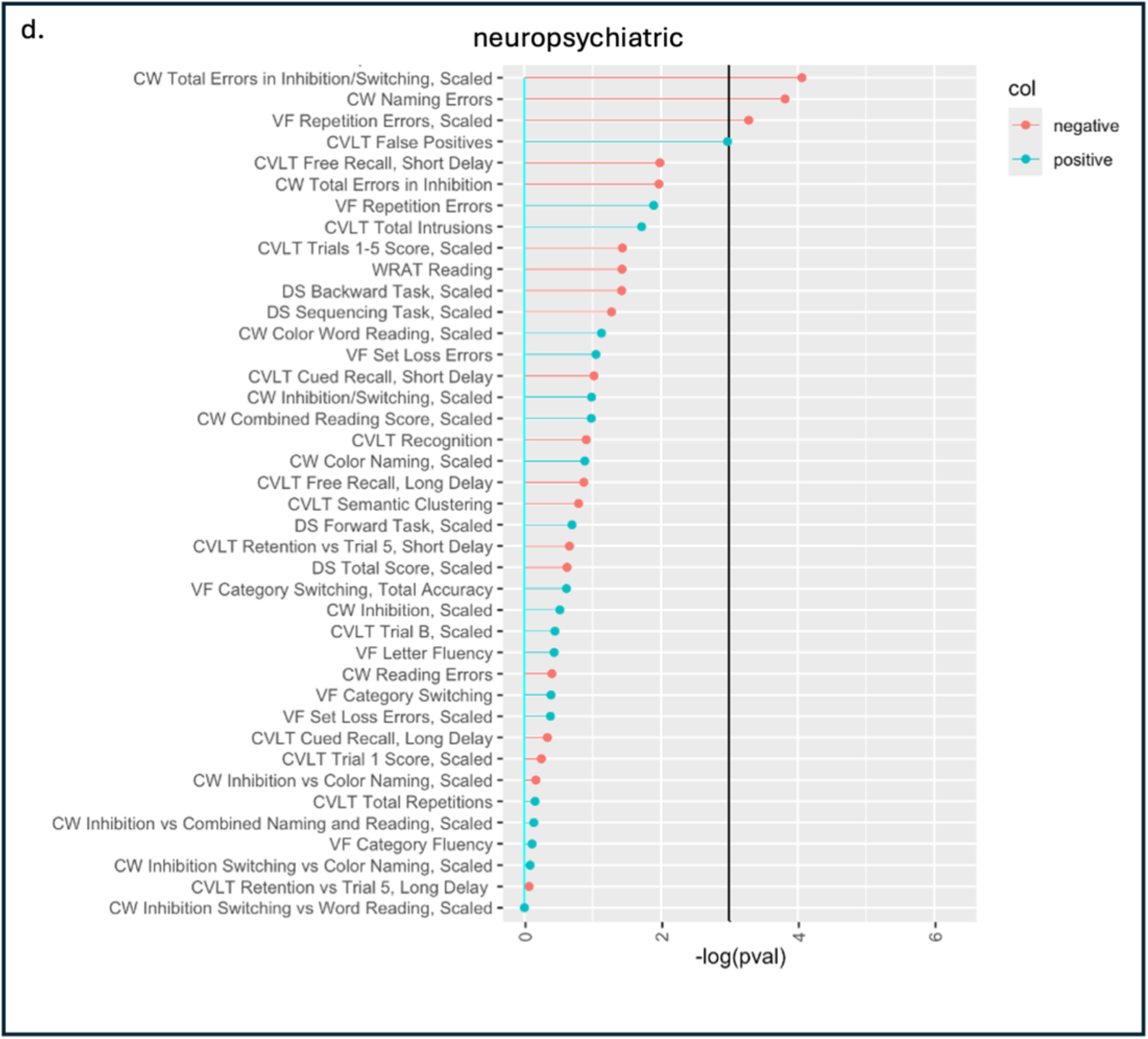
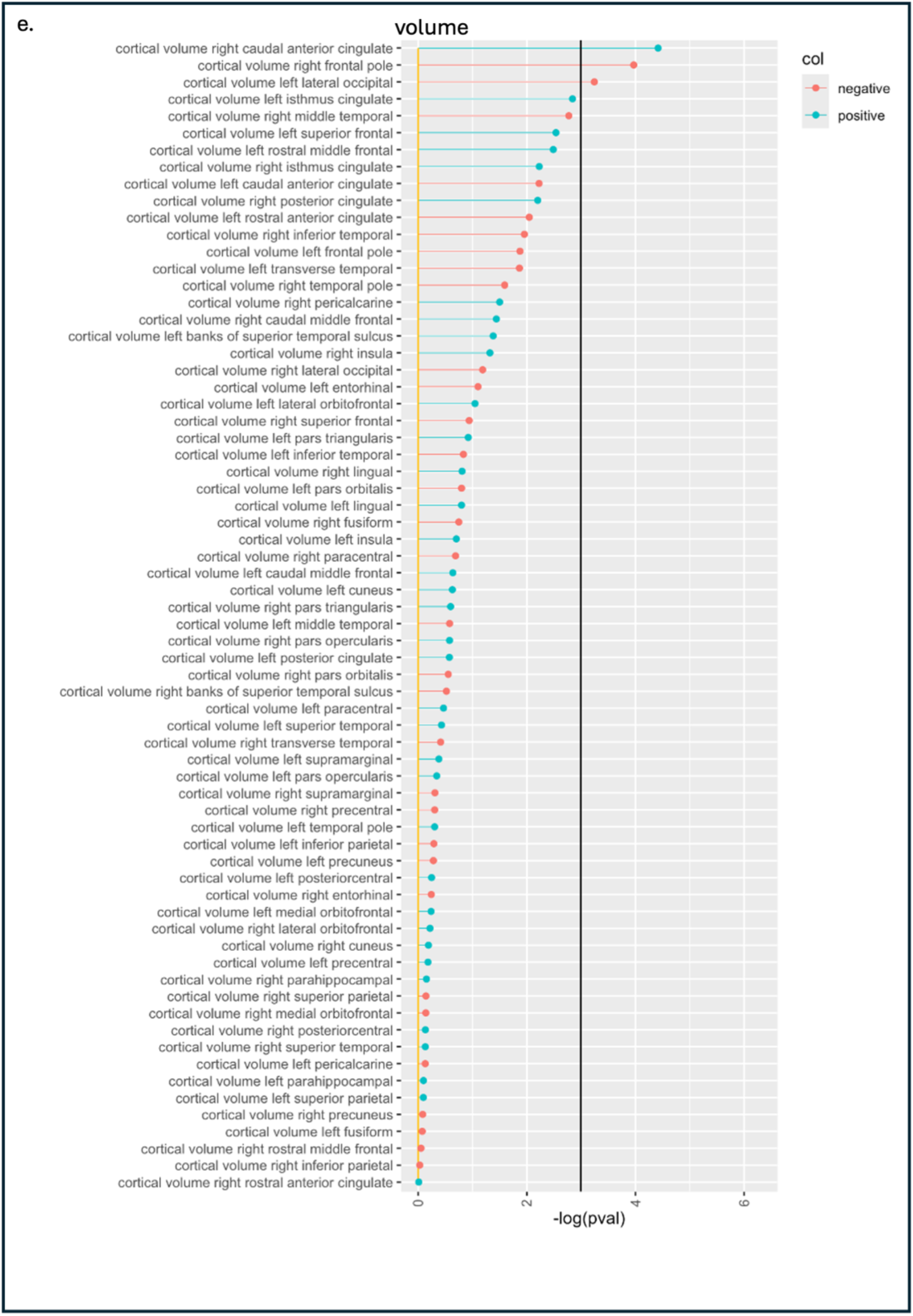
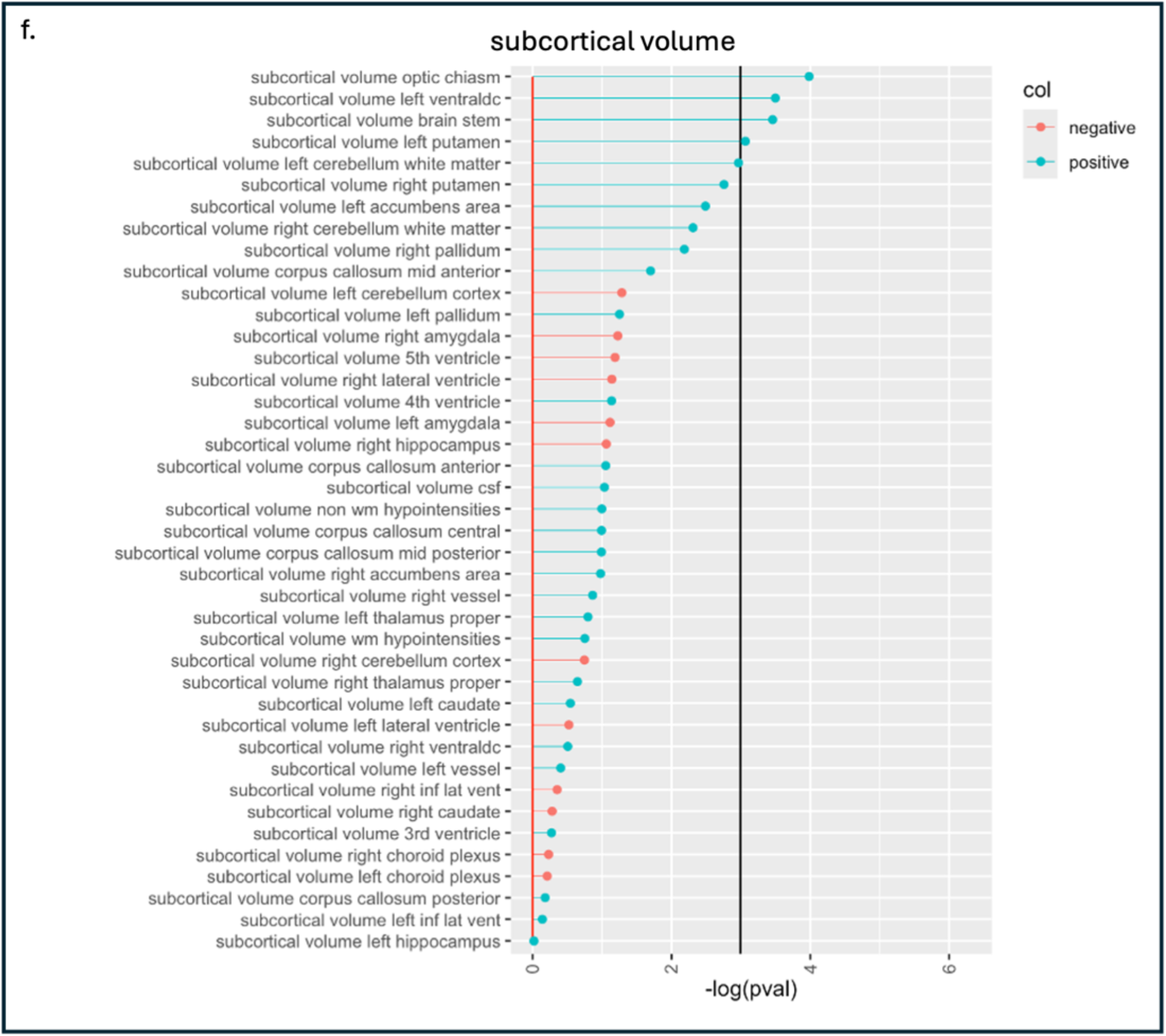
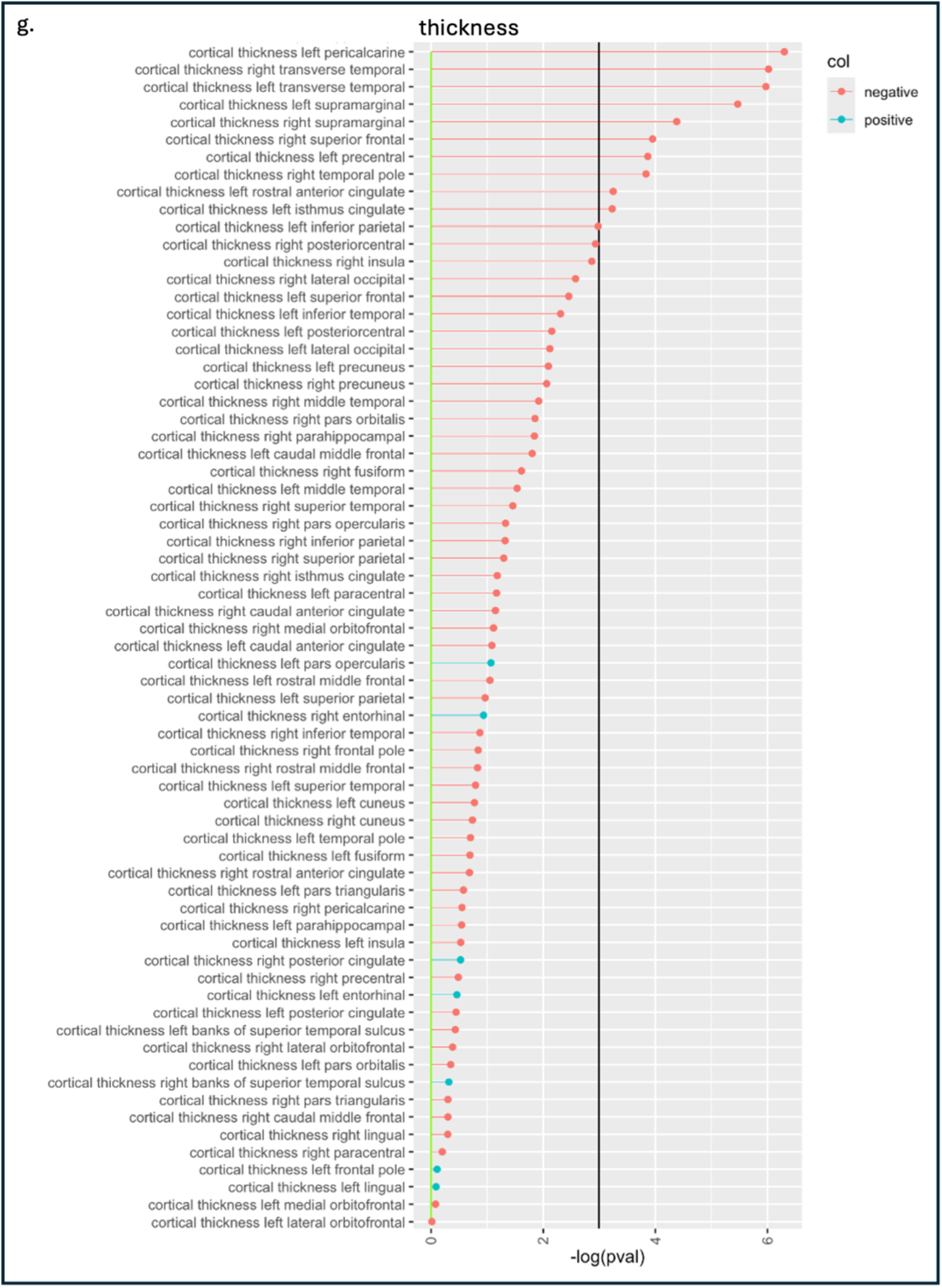
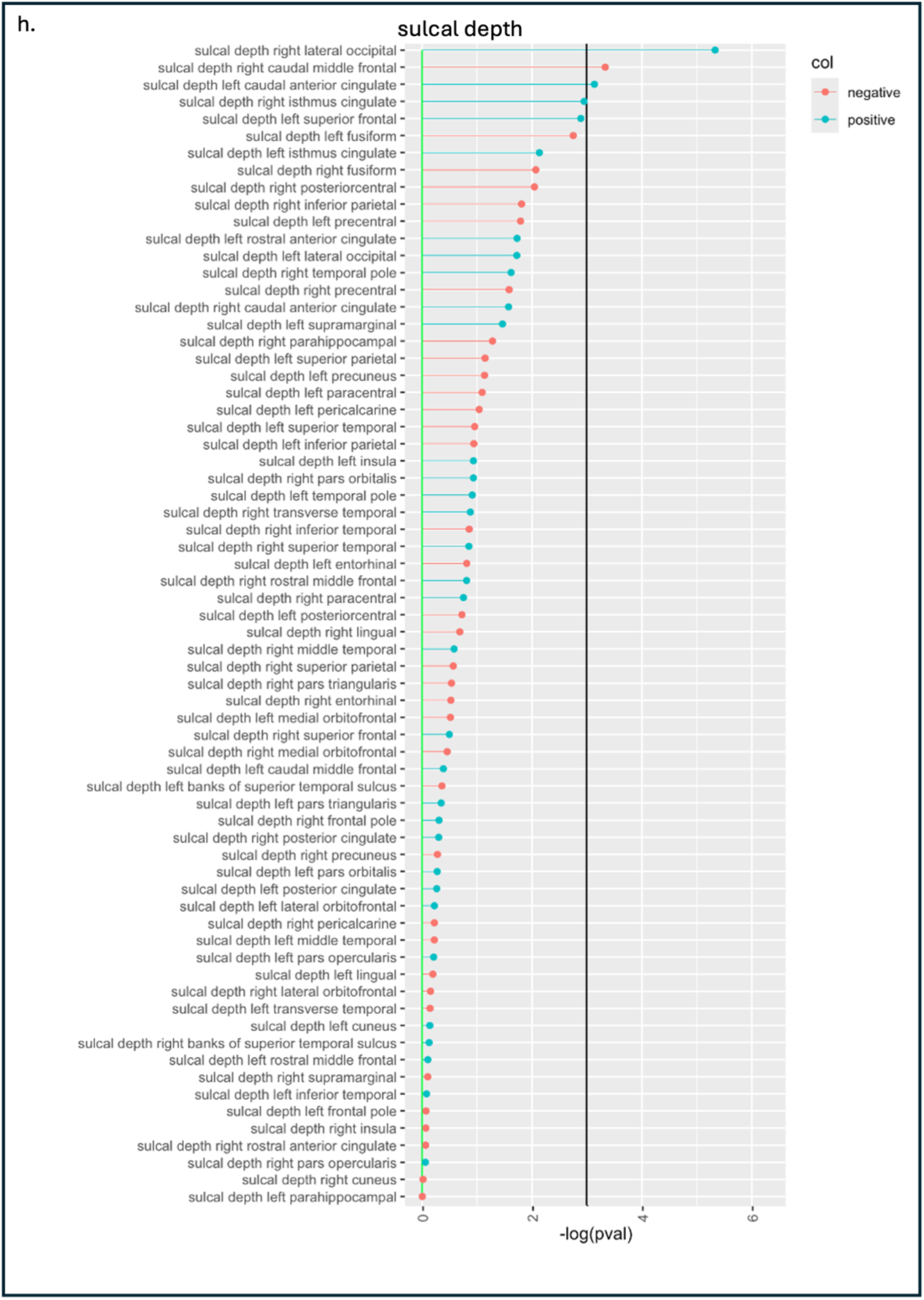
Each plot depicts the negative log of the p-value of the association between risk allele count and the predictor variables when controlling for age, gender, education, BMI, and cortical volume. Blue dots indicate a positive relationship and red points indicate a negative relationship. The black line marks p < 0.05. Plots are separated by predictor type: (a) drug, (b) diet, (c) symptom, (d) neuropsychiatric, (e) volume, (f) subcortical volume, (g) thickness, (h) sulcal depth.

‘Eats Eggs’ was the strongest predictor of risk allele count and this relationship was statistically significant, with egg consumption being positively related to risk alleles. Vegetarianism was the next strongest predictor, though not significant, with vegetarianism associated with lower risk allele count. Analgesic use was also significantly predictive of risk allele count, with analgesic use related to lower risk allele count (Figure 2.a.). Some structural variables were indicated as having the largest variable importance, including cortical thickness left pericalcarine, cortical thickness right transverse temporal, cortical thickness left transverse temporal, cortical thickness left supramarginal, and sulcal depth right lateral occipital (Figure 2.b.)

**FIGURE 2.A.**
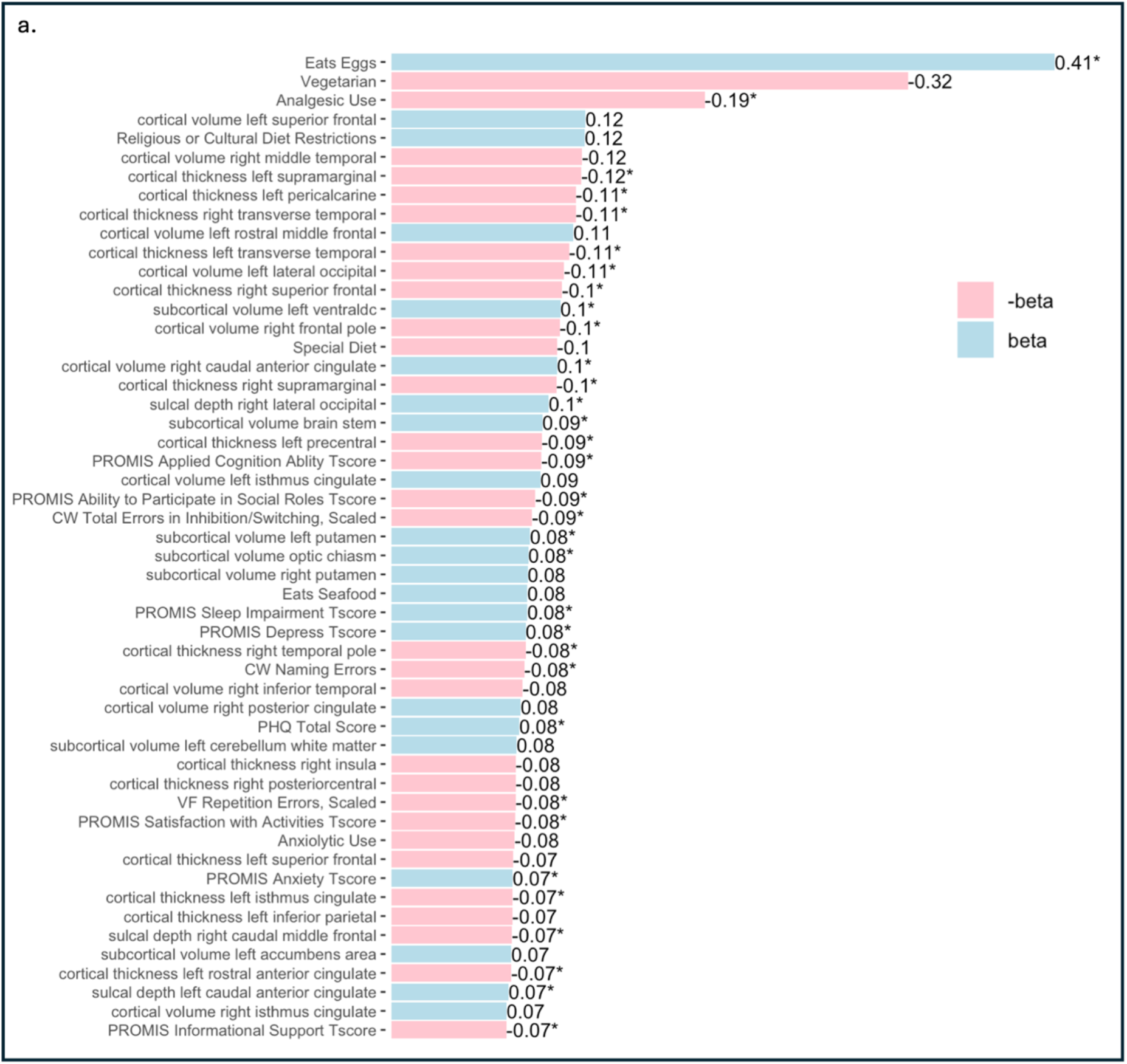
This plot depicts the highest-magnitude beta values from the linear regressions predicting risk allele count and covarying for age, gender, education, BMI, and cortical volume. Asterisks mark beta values with a p-value < 0.05. The smallest-magnitude beta value that maintained statistical significance was PROMIS Informational Support T-Score with a beta of −0.07, and variables with lower-magnitude betas were excluded from the plot for the sake of readability.

**FIGURE 2.B.**
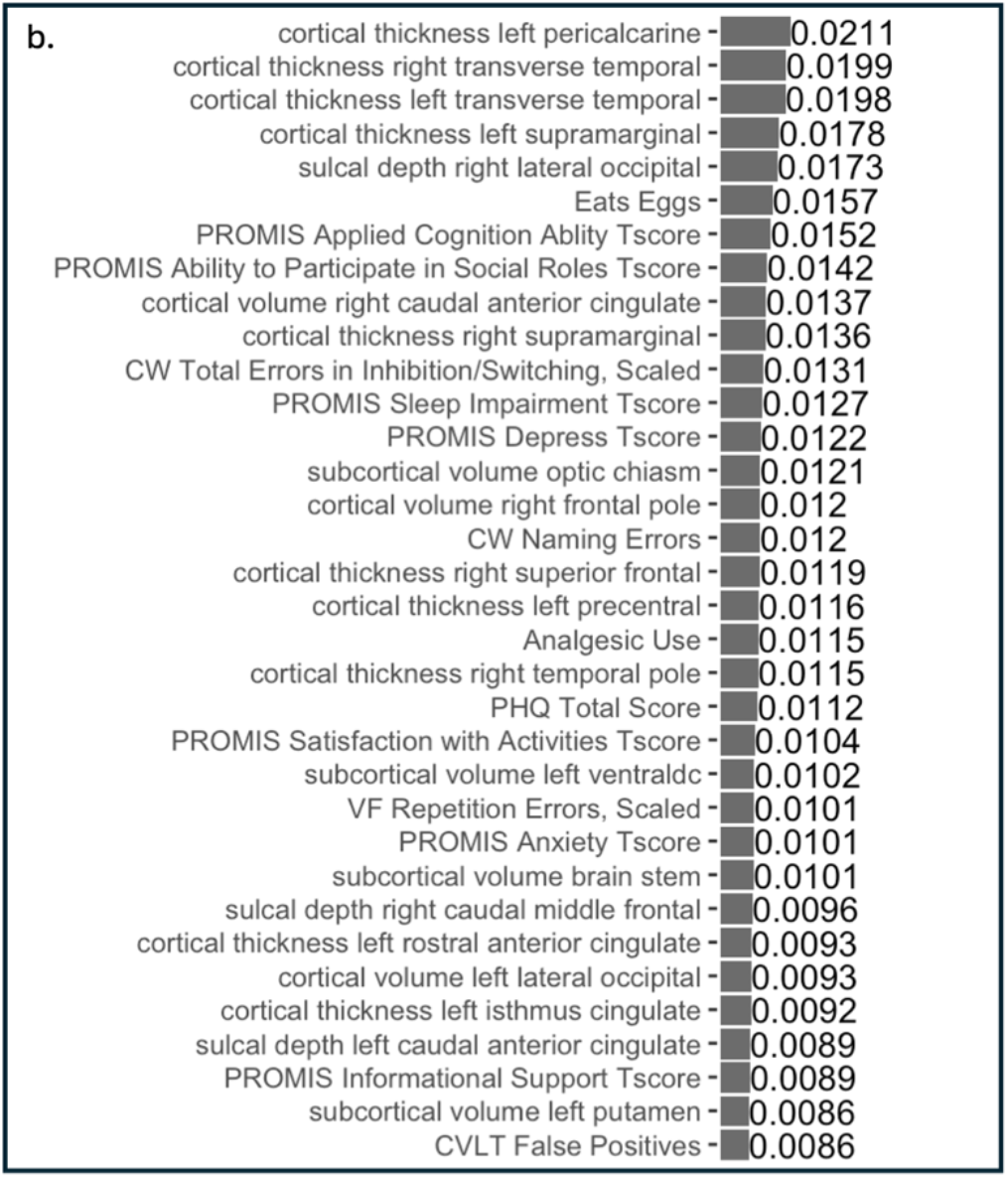
This plot depicts the largest variable importance, calculated as the R-square differences between the models with and without the indicated predictor variable. All predictors with a variable importance of 0.0086 or higher were statistically significant. Predictors with a variable importance below 0.0086 were not statistically significant and were not included in the plot for the sake of readability.

**TABLE 1.**
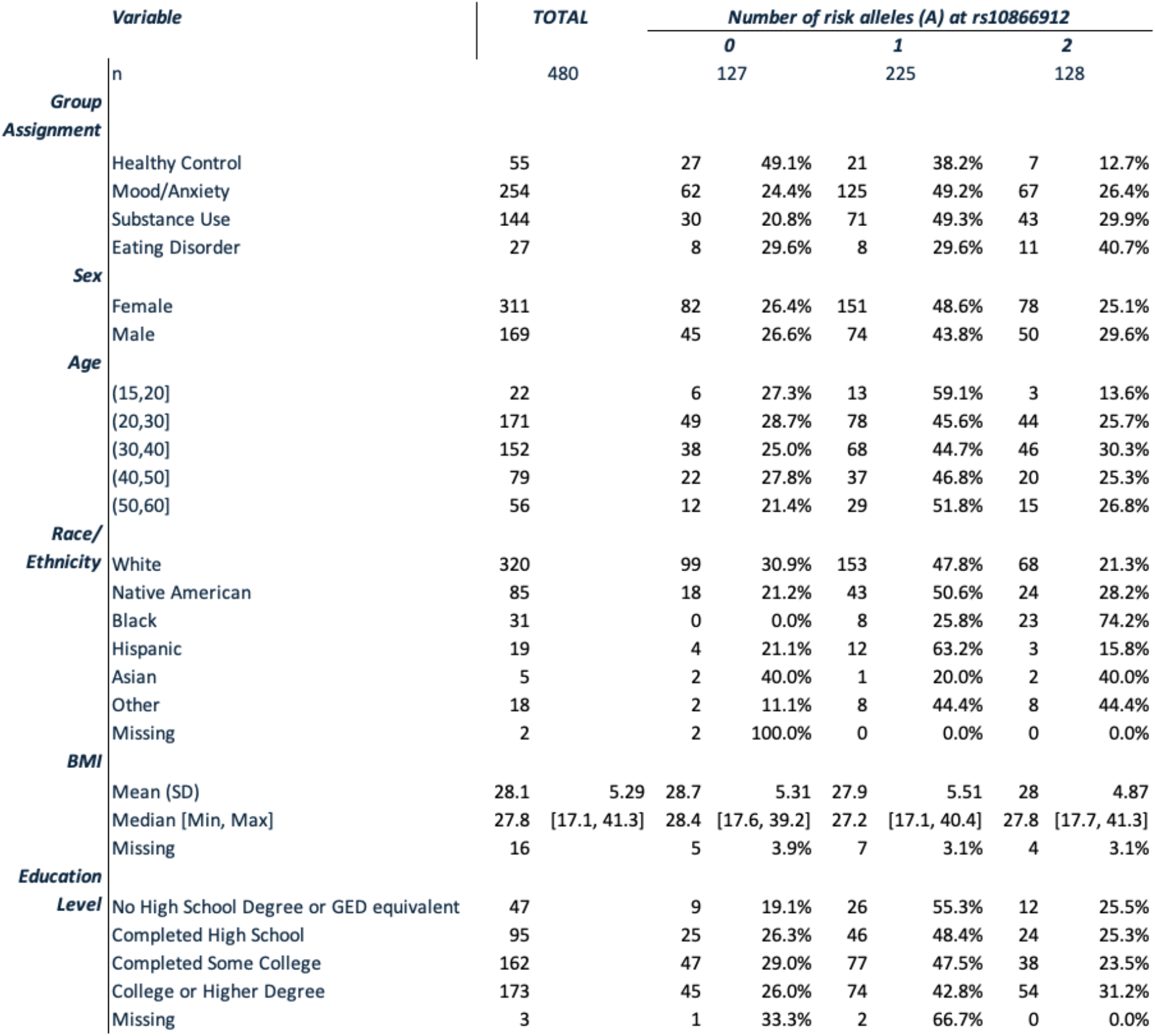
In this table, we compare the demographic distribution within each risk allele count.

## Discussion

From our study of 500 participants, we identified the SNP rs10866912 in the imputed genome sequence of 480 individuals. Within our sample, 47% had one copy of the allele, 27% had two copies, and 26% had no copies. Notably, healthy controls predominantly lacked the risk allele, whereas individuals with mood/anxiety disorders, substance use disorders, and eating disorders more frequently carried one or two copies of the risk allele. Our linear models identified 33 predictor variables significantly associated with risk allele count, though none survived corrections for multiple comparisons. Significant associations included various symptom measures, neuropsychiatric test results, diet, drug use, and cortical structures. Notably, egg consumption was the strongest predictor of risk allele count, with egg consumption positively associated with the number of risk alleles.

The SNP rs10866912 has been implicated in various health conditions through genome-wide association studies (GWAS). This SNP, located in a region likely involved in regulatory functions, influences gene expression and potentially contributes to disease susceptibility. Our preliminary results align with previous research indicating that prevalence of the risk allele varies between demographic and clinical populations. Within our sample, individuals with mood and anxiety, substance use, and eating disorders exhibited a higher prevalence of the risk (A) allele than the healthy population (Psychiatric Disorders: 23.5% with 0 alleles, 48% with 1 allele, and 28.5% with 2 alleles | Healthy Controls: 49.1% with 0 alleles, 38.2% with 1 allele, and 12.7% with 2 alleles). This suggests that rs10866912 may be linked to neurobiological mechanisms underlying psychiatric disorders. The risk allele’s prevalence in individuals with substance use disorders is of particular note as this population has the largest share of individuals with at least one copy of the risk allele (20.8% with 0 alleles, 49.3% with 1 allele, and 29.9% with 2 alleles), highlighting its potential role in addiction pathways. The stark racial/ethnic differences underscore the impact of genetic ancestry on the population prevalence of the risk allele. Our findings align with previous research that indicates individuals of African descent as having particularly high risk of inheriting the risk allele (12). This points to a need for inclusive genetic research to understand the effects of this allele more clearly.

Our primary analysis identified significant linear associations between the frequency of the risk allele at rs10866912 and measures describing drug use, diet, psychiatric symptoms, neuropsychiatric ability, and structural brain measures including cortical and subcortical volumes, cortical thickness, and sulcal depth, though no associations survived correction for multiple comparisons.

Of the drug variables, analgesic use significantly predicted the risk allele, with analgesic use being associated with absence of the risk allele. Of the diet variables, egg consumption was another significant predictor, with egg consumption positively associated with presence of the risk allele. We speculate that individuals homozygous for the risk allele have a downregulated Preiss-Handler pathway and thus may be more likely to consume niacin-rich food groups such as eggs in an attempt to upregulate that pathway by providing additional dietary niacin.

Among the psychiatric symptom variables, PHQ total score and PROMIS depression and anxiety t-scores were indicated as positively predicting the risk allele. Other significant symptom variables indicated measures on cognitive ability and social satisfaction to be negatively associated with the risk allele, and sleep impairment to be positively associated. Significant results among the neuropsychiatric measures indicated a negative relationship between cognitive abilities and presence of the risk allele. Overall, the results relating psychiatric symptoms and neuropsychiatric tests to the risk allele were consistent, with the risk allele being broadly associated with poor psychiatric outcomes and reduced cognitive abilities.

Several of the structural measures were also of note. Areas of the brain where volume or sulcal depth was positively associated with the risk allele included the optic chiasm (subcortical volume; scv), left ventral diencephalon (scv), right caudal anterior cingulate (cortical volume; cv), left caudal anterior cingulate (sulcal depth; sd), brain stem (scv), left putamen (scv), and right lateral occipital (sd). These regions are functionally related to vision (optic chiasm), motor control (putamen), emotional processing (anterior cingulate), and basic physiological processes (brain stem). Areas of the brain where volume or sulcal depth were negatively associated with the risk allele included the right frontal pole (cv), right caudal middle frontal (sd), and left lateral occipital (cv). These regions are associated with cognitive function (frontal regions) and visual processing (occipital region). All significant associations among the cortical thickness measures indicated a negative relationship with the risk allele, with particular significance indicated within right superior frontal, left precentral, left rostral anterior cingulate, left transverse temporal, right transverse temporal, right temporal pole, left supramarginal, right supramarginal, left isthmus cingulate, and left pericalcarine. These relationships indicate that presence of the risk allele is associated with reduced cortical thickness (13) in regions involved in a range of functions including executive function (frontal regions), motor control (precentral), emotional regulation (anterior cingulate), auditory processing (transverse temporal), and sensory integration (supramarginal). These findings indicate a complex interplay between genetic factors and brain morphology, resonating with findings from neuroimaging genetics.

Several limitations should be considered when interpreting these results. First, our linear regressions did not incorporate genetic ancestry as a covariate. As the risk allele is strongly related to genetic ancestry, this could suggest that significant results are confounded by genetic ancestry. Second, the absence of significant findings after correction for multiple comparisons suggests that significant results could be due to a Type 1 error, indicating the need for larger sample sizes to validate these findings.

## Conclusion

The SNP rs10866912 represents a significant genetic marker for understanding the etiology of complex diseases, including psychiatric disorders. This study indicates that the risk (A) allele is more common amongst individuals with psychiatric disorders. Mechanisms may be implied by the relationships indicated with our linear models, in particular relationship between the risk allele and egg consumption, analgesic use, cognitive abilities, psychiatric symptoms, and cortical structure. Despite the limitations, these results contribute to our understanding of the genetic factors influencing mental health. Future research should focus on the functional implications of rs10866912 and its role in gene regulation, which could pave the way for novel therapeutic strategies.

## Data Availability

All data produced in the present study are available upon reasonable request to the authors

## Acknowledgements

This work has been supported in part by the William K. Warren Foundation and The University of Queensland, Australia. The funders had no role in study design, in the collection, analysis, and interpretation of data, in the writing of the manuscript, or in the decision to submit the paper for publication.

Participants provided written informed consent and received remuneration for study participation according to the study protocol approved by the Western-Copernicus Group Institutional Review Board (IRB Tracking Number: 20142082).

The ClinicalTrials.gov identifier for the clinical protocol associated with data published in the current paper is NCT02450240, “Latent Structure of Multi-level Assessments and Predictors of Outcomes in Psychiatric Disorders” (https://clinicaltrials.gov/ct2/show/NCT02450240).

Drs. Paulus, Mowry, Periyasamy, and Ms. Forthman have no competing interests to disclose.

